# Living alone, loneliness and lack of emotional support as predictors of suicide and self-harm: a nine-year follow up of the UK Biobank cohort

**DOI:** 10.1101/19008458

**Authors:** Richard J. Shaw, Breda Cullen, Nicholas Graham, Donald M. Lyall, Daniel Mackay, Chukwudi Okolie, Robert Pearsall, Joey Ward, Ann John, Daniel J. Smith

## Abstract

**Background:** The association between loneliness and suicide is poorly understood. We investigated how living alone, loneliness and emotional support were related to suicide and self-harm in a longitudinal design.

**Methods:** Between 2006 and 2010 UK Biobank recruited and assessed in detail over 0.5 million people in middle age. Data were linked to prospective hospital admission and mortality records. Adjusted Cox regression models were used to investigate relationships between living arrangements, loneliness and emotional support, and both suicide and self-harm as outcomes.

**Results:** For men, both living alone (Hazard Ratio (HR) 2.16, 95%CI 1.51-3.09) and living with non-partners (HR 1.80, 95%CI 1.08-3.00) were associated with death by suicide, independently of loneliness, which had a modest relationship with suicide (HR 1.43, 95%CI 0.1.01-2.03). For women, there was no evidence that living arrangements, loneliness or emotional support were associated with death by suicide. Associations between living alone and self-harm were explained by health for women, and by health, loneliness and emotional support for men. In fully adjusted models, loneliness was associated with hospital admissions for self-harm in both women (HR 1.89, 95%CI 1.57-2.28) and men (HR 1.74, 95%CI 1.40-2.16).

**Limitations:** Loneliness and emotional support were operationalized using single item measures.

**Conclusions:** For men - but not for women - living alone or living with a non-partner increased the risk of suicide, a finding not explained by subjective loneliness. Overall, loneliness may be more important as a risk factor for self-harm than for suicide. Loneliness also appears to lessen the protective associations of cohabitation.

**Highlights:**

- First cohort study to investigate loneliness’s relationship with deaths by suicide
- Loneliness is associated with a modest increased risk of death by suicide for men
- For men, living with a partner reduces the risk of death by suicide
- Loneliness increases the risk of hospitalization for self-harm for men and women

## INTRODUCTION

Loneliness, defined as *the subjective perception of a lack of contact with other people* (HM Government, 2018; Perlman and Peplau, 1981), is associated with premature mortality (Elovainio et al., 2017; Rico-Uribe et al., 2018), physical and mental ill-health, worse cognitive function (Cacioppo et al., 2014; Hakulinen et al., 2018; Hawkley and Cacioppo, 2010; Solmi et al., 2020) and increased use of health services (Dreyer et al., 2018). Loneliness affects people of all ages (Age UK, 2018) and has been made a ministerial responsibility by the UK government (HM Government, 2018). While living alone has been consistently linked with self-harm and suicide, it is currently not clear whether subjective loneliness *per se* is the primary reason why people living alone may be at increased risk of suicidal behaviour.

Living alone and loneliness indicate relationships and social connections, nonetheless they are separate constructs with overlapping features (Smith and Victor, 2019). Living alone is distinct from cohabitating relationships, as well as residency with non-partners such as parents, children or friends, who might be expected to be sources of emotional, financial and practical support (Amato, 2014; van Hedel et al., 2018). However, clearly people living alone may engage with others outside the household and potentially receive emotional support from other sources. In contrast, loneliness is the *subjective perception* of a lack of contact with other people (Hawkley and Cacioppo, 2010). Emotional support is a related concept. It involves the provision of caring, empathy, love and trust within a relationship (Langford et al., 1997) and indicates that somebody is taken care of, valued, not alone and has somebody to confide in (Shensa et al., 2020; Yao et al., 2015).

Theories that would support a relationship between suicide and loneliness, living arrangements and lack of emotional support, trace back to the work of Durkheim (Stanley et al., 2016). In particular, egoistic suicide is described as a lack of social integration because of reasons such as an individual’s lack of social bonds to family and friends. These ideas have been developed further in modern theories such as the Interpersonal Theory of Suicide. One aspect of the Interpersonal Theory of Suicide of particular relevance is the concept of ‘thwarted belongingness’ which suggests that loneliness along with the absence emotional support can lead to self-destructive behaviours (Stanley et al., 2016; Van Orden et al., 2010). However, there is little empirical research in this area (Van Orden et al., 2010). Identifying robust risk factors for suicidal behaviour is methodologically challenging for a number of reasons, not least because suicide is a rare event (Klonsky et al., 2016; Stickley and Koyanagi, 2016). Most studies of loneliness and suicidal behaviour have used self-reported measures of suicidality (Bennardi et al., 2019), which may be prone to reporting biases (Beutel et al., 2017; Stickley and Koyanagi, 2016), and only a few studies (mostly case-control studies) have investigated loneliness as a potential cause of deaths by suicide (Courtin and Knapp, 2017; Holt-Lunstad et al., 2015).

The extensive data within the UK Biobank cohort represents a unique opportunity to overcome these methodological challenges. The cohort consists of over half a million people, and the baseline questionnaire included detailed questions on living arrangements, loneliness and emotional support, in addition to key sociodemographic and health data. These data have also been linked prospectively to hospital episode statistics and mortality records (Sudlow et al., 2015).

Our primary hypothesis was that living alone may represent an independent risk factor for self-harm and suicide. We also set out to assess whether any observed association between living alone and suicidal behaviour might be explained by subjective loneliness or by perceived lack of emotional support.

## METHODS

### Data

All adults aged between 40-70 years who were registered with the UK National Health Service (NHS) and living within 25 miles of 22 assessment centres across England, Scotland and Wales were invited to participate in UK Biobank at baseline. The achieved sample of 502,536 people had a response rate of 5.5% (Sudlow et al., 2015) and an age range of between 37 and 73. Participants were recruited between March 2006 and October 2010 (Sudlow et al., 2015) and for each participant baseline assessments consisted of a single visit lasting approximately two to three hours, including a computerised self-completion touch screen questionnaire, nurse interviews, and physical measurements. For all participants, the date and cause of death were sought from death certificates held within the National Health Service Information Centre (England and Wales), and National Health Service Central Register Scotland. At the time of analysis, mortality data were available up to the middle of February 2018 for England and Wales and until June 2017 for Scotland. Data for Hospital admission records for self-harm were only available for participants in England and for the period up to March 2015, thus analyses for self-harm were restricted to those who attended a Biobank assessment centre in England (n = 448,811). This study is covered by the generic ethical approval for UK Biobank studies from the National Health Service National Research Ethics Service (June 17, 2011; Ref 11/NW/0382). Participants provided electronic informed consent for the baseline assessments and the register linkage.

### Outcomes

*Death by suicide* was defined as the act of intentionally ending one’s own life (Nock et al., 2008) and was ascertained from death records using ICD 10 codes X60-X84 (intentional self-harm), Y10-34 (undetermined cause), as used by the UK Office for National Statistics (2019). Participants dying from other causes of death were censored at time of death.

*Hospital admissions for self-harm* were defined as any act of intentional self-poisoning or self-injury carried out by an individual, irrespective of the motivation or suicidal intent (National Collaborating Centre for Mental Health, 2012). This was assessed using the first admission for self-harm following attendance at the UK Biobank baseline assessment centre. Hospitalization for self-harm was assessed using ICD 10 codes X60 to-84 and Z91.5, for diagnosis and causes of admissions.

### Main exposures of interest

*Living arrangements* (alone; husband, wife or partner; other) were assessed using data from the baseline touch screen questionnaire collected when participants first attended a UK Biobank assessment centre. Participants were asked how many people lived in their household. If there was more than one person, the participant was then asked how people were related to them. If any member of the household was a spouse or partner, participants were classified as living with a husband, wife or partner. The other category included both relatives and unrelated people.

*Loneliness* was assessed using a single question taken from the baseline touchscreen questionnaire: “Do you often feel lonely?” (Responses: yes; no; do not know; prefer not to answer). This item was taken from a longer scale and has previously been shown to be associated with health outcomes (McCormack et al., 2014).

*Emotional Support* was assessed with the question “How often are you able to confide in someone close to you?” (Shensa et al., 2020). The potential responses were “Almost daily; 2-4 times a week; about once a week; about once a month or once every few months; never or almost never; do not know or prefer not answer” (the latter were coded as missing).

### Covariates

We included sociodemographic and health variables which, which were collected during participants’ attendance at baseline assessment centres, and might confound relationships between the key variables of interest and death by suicide and hospital admissions for self-harm.

The socio-demographic variables included were: sex, age (continuous), and self-reported measures of ethnicity (derived here into White British, other), current employment status, (employed, retired, other), education (degree; professional; NVQ, HND or HNC; A level; O level; CSE; none) ever having a same sex partner (none, at least one), and area deprivation indicated using the Townsend Index (continuous) for the participant’s postcode at recruitment.

The health variables included were: a measure of multimorbidity, developed for a previous study (Nicholl et al., 2014) which was the number of physical morbidities participants reported to the interviewing nurse (zero, one, two, three or more); Body Mass Index (BMI) based on measurements made at the assessment centre (normal or underweight, overweight, obese); self-reported measure of depression based on of ever seeing a GP for nerves, anxiety tension or depression (yes, no); participants’ report during their baseline interview of taking psychotropic medication (yes/no); alcohol consumption (daily/almost daily, 3-4 days a week, 1-2 times a week /once a month, special occasions/never, former); and smoking status (never, previous, current).

### Statistical analysis

Prior research (Kyung-Sook et al., 2018), and based on statistical interactions we found in preliminary analyses, indicated that gender modifies the relationship between the main exposures of interest and suicidality. Consequently, we carried out the analyses stratified by gender. Cox proportional hazards regression was used to investigate deaths by suicide. The proportional hazards assumption was tested for using Schoenfeld residuals. For hospital admissions due to self-harm, however, the proportional hazards assumption was not met for loneliness, so data were reanalysed using a Royston Parmar model (Royston and Lambert, 2011), with Akaike information criterion from preliminary analyses indicating that two knots should be used to model the baseline hazard, and single time varying parameter, for loneliness.

For each participant the start date for the follow-up period used in analyses was the date of their first attendance at a UK Biobank centre at baseline, which ranged from March 2006 to October 2010. Participants were censored upon death, and for the death by suicide analyses, the last date (February 2018 for England and Wales and until June 2017 for Scotland) that mortality records were available, and for the analyses of self-harm, the last date (March 2015) that hospital records were available.

Six different models are presented for both deaths by suicide and hospital admissions for self-harm. The first three models are presented for each of living arrangements, loneliness, and emotional support separately. Models 1 are univarable regression models only including each of the main independent variables. Models 2 adjust for all sociodemographic variables, and Models 3 additionally adjust for the health variables. Model 4 adds loneliness and Model 5 adds emotional support to Model 3. In Model 6, all variables were included.

We accounted for missing data using multiple imputation by chained equations, generating twenty imputed data sets. Imputation models were stratified by gender and included age, living arrangements, loneliness, emotional support, all variables used in the models, the Nelson-Aalen estimate of cumulative hazard, survival status (Cleves et al., 2016), and additional variables to improve model fit, including household income, participation in social groups, contact with friends and family, parental depression, limiting longstanding illness and self-rated health. These variables were not included in the main models because they either had comparatively high rates of missing data which limited their utility in preliminary complete case analysis, or, in the case of health variables, might mediate the relationship between our exposures of interest and outcomes. Our models were fitted to each imputed data set and combined in accordance with Rubin’s rules. All analyses were carried out using Stata 16.0.

## RESULTS

Sociodemographic characteristics of study participants by gender are shown in table 1. With respect to the main independent variables of interest, men were more likely to cohabit, whereas women were more likely to live alone or with non-partner(s). Women were somewhat more likely to report often feeling lonely, and men were much more likely to report that they never had any emotional support. Men were much more likely to have died by suicide (n=181, 8.9 deaths per 100,000 participants per year) than women (n=85, 3.5 deaths per 100,000 participants per year). Among potential confounders, men were more likely than woman to be in both the most and least advantaged categories of the socioeconomic measures, and women generally had poorer health.

**Table 1:** Characteristics for Men and Women in UK Biobank.

|  | Women |  | Men |  |  | Women |  | Men |  |
| --- | --- | --- | --- | --- | --- | --- | --- | --- | --- |
|  | n | % | n | % |  | n | % | n | % |
| *Social Connection* |  |  |  |  |  |  |  |  |  |
| **Living arrange<sup>\*</sup>** |  |  |  |  | **Centre Country** |  |  |  |  |
| Alone | 53,737 | 19.7 | 39,188 | 17.1 | England | 243,756 | 89.2 | 205,057 | 89.5 |
| Cohabitation | 188,588 | 69.0 | 174,503 | 76.2 | Wales | 9,701 | 3.6 | 8,177 | 3.6 |
| Non Partner | 28,787 | 10.5 | 13,330 | 5.8 | Scotland | 19,945 | 7.3 | 15,900 | 6.9 |
| Missing | 2,290 | 0.8 | 2,113 | 0.9 |  |  |  |  |  |
| **loneliness** |  |  |  |  | *Health* |  |  |  |  |
| No | 210,917 | 77.2 | 190,330 | 83.1 | **Multimorbidity<sup>1</sup>** |  |  |  |  |
| Yes | 57,045 | 20.9 | 34,371 | 15.0 | Zero | 99,438 | 36.4 | 83,795 | 36.6 |
| Missing | 5,440 | 2.0 | 4,433 | 1.9 | One | 90,116 | 33.0 | 76,252 | 33.3 |
| **Emotional Support** |  |  |  |  | Two | 49,995 | 18.3 | 42,382 | 18.5 |
| Almost Daily | 140,692 | 51.5 | 117,881 | 51.5 | Three or more | 33,853 | 12.4 | 26,705 | 11.7 |
| 2-4 times a week | 30,321 | 11.1 | 16,462 | 7.2 | missing | na<sup>1</sup> |  | Na |  |
| Once a Week | 33,579 | 12.3 | 19,751 | 8.6 | **Ever Depressed** |  |  |  |  |
| Monthly | 30,276 | 11.1 | 22,965 | 10.0 | No | 159,662 | 58.4 | 168,388 | 73.5 |
| Never | 29,284 | 10.7 | 42,449 | 18.5 | Yes | 110,878 | 40.6 | 58,590 | 25.6 |
| Missing | 9,250 | 3.4 | 9,626 | 4.2 | Missing | 2,862 | 1.1 | 2,156 | 0.9 |
| *Socio Demographic* |  |  |  |  | **Psychotropic Meds** |  |  |  |  |
| **Age** |  |  |  |  | No | 235,483 | 86.1 | 209,628 | 91.5 |
| 35/44 | 27,835 | 10.2 | 23,968 | 10.5 | Yes | 28,474 | 10.4 | 14,217 | 6.2 |
| 45/54 | 80,110 | 29.3 | 62,305 | 27.2 | Missing | 9,445 | 3.5 | 5,289 | 2.3 |
| 55/64 | 116,860 | 42.7 | 95,446 | 41.7 | **Alcohol usage** |  |  |  |  |
| 65/74 | 48,597 | 17.8 | 47,415 | 20.7 | Daily / almost daily | 43,867 | 16.0 | 57,907 | 25.3 |
| **Ethnicity** |  |  |  |  | 3 to 4 times week | 55,899 | 20.5 | 59,546 | 26.0 |
| White British | 240,265 | 87.9 | 202,345 | 88.3 | 1 -2 times a week | 105,686 | 38.7 | 79,469 | 34.7 |
| Other | 31,871 | 11.7 | 25,278 | 11.0 | Special event or never | 57,137 | 20.9 | 23,263 | 10.2 |
| Missing | 1,266 | 0.5 | 1,511 | 0.7 | Former | 9,983 | 3.7 | 8,125 | 3.6 |
| **Employment status** |  |  |  |  | Missing | 830 | 0.3 | 824 | 0.4 |
| Employed | 149,212 | 54.6 | 137,958 | 60.2 | **Smoking** |  |  |  |  |
| Retired | 95,565 | 35.0 | 71,420 | 31.2 | Never | 162,064 | 59.3 | 111,473 | 48.7 |
| Other | 27,105 | 9.9 | 18,323 | 8.0 | Previous | 85,458 | 31.3 | 87,612 | 38.2 |
| Missing | 1,520 | 0.6 | 1,433 | 0.6 | Current | 24,367 | 8.9 | 28,612 | 12.5 |
| **Deprivation** |  |  |  |  | Missing | 1,513 | 0.6 | 1,437 | 0.6 |
| Least deprived | 54,571 | 20.0 | 46,093 | 20.1 | **Obesity** |  |  |  |  |
| 4<sup>th</sup> Quintile | 54,568 | 20.0 | 45,537 | 19.9 | Normal & Under | 107,822 | 39.4 | 57,340 | 25.0 |
| 3<sup>rd</sup> Quintile | 55,333 | 20.2 | 45,057 | 19.7 | Overweight | 99,859 | 36.5 | 112,249 | 49.0 |
| 2<sup>nd</sup> Quintile | 55,355 | 20.3 | 45,020 | 19.7 | Obese | 64,262 | 23.5 | 57,899 | 25.3 |
| Most deprived | 53,248 | 19.5 | 47,131 | 20.6 | Missing | 1,459 | 0.5 | 1,646 | 0.7 |
| Missing | 327 | 0.1 | 296 | 0.1 | *Suicidality* |  |  |  |  |
| **Education** |  |  |  |  | **Hospital admission for** |  |  |  |  |
| Degree | 84,541 | 30.9 | 76,635 | 33.4 | **Self-harm<sup>2</sup>** |  |  |  |  |
| Professional | 40,982 | 15.0 | 31,452 | 13.7 | N Events | 589 |  | 471 |  |
| NVQ, HND or HNC | 28,304 | 10.4 | 34,682 | 15.1 | Median time to event (yrs) | 2.37 |  | 2.61s |  |
| A level | 15,758 | 5.8 | 11,477 | 5.0 | N Censored due to death | 3,966 |  | 6,173 |  |
| O level | 41,639 | 15.2 | 23,045 | 10.1 | Median time to censor (yrs) | 3.77 |  | 3.64 |  |
| CSE | 10,863 | 4.0 | 7,745 | 3.4 | Person yrs follow up | 1,458,045 |  | 1,220,987 |  |
| None | 45,968 | 16.8 | 39,307 | 17.2 | **Died by suicide** |  |  |  |  |
| Missing | 5,347 | 2.0 | 4,791 | 2.1 | N Events | 85 |  | 181 |  |
| **Ever same sex** |  |  |  |  | Median time to event (yrs) | 5.00 |  | 4.54 |  |
| **Relationship** |  |  |  |  | N Censored due death | 7,981 |  | 12,037 |  |
| None | 235,296 | 86.1 | 197,726 | 86.3 | Median time to censor (yrs) | 5.64 |  | 5.54 |  |
| At least one | 6,923 | 2.5 | 8,894 | 3.9 | Person yrs follow up | 2,444,031 |  | 2,027,824 |  |
| Missing | 31,184 | 11.4 | 22,514 | 9.8 |  |  |  |  |  |

1. Data are only available for those reporting conditions,
2 Analyses only include those who attended a Biobank centre in England at baseline.

### Death by suicide

The results of Cox proportional hazard models for deaths by suicide are shown in table 2. For men there were initially strong relationships, relative to cohabitation, between living alone or with a non-partner and death by suicide (model 1). These associations were attenuated after adjusting for sociodemographic factors (sex, age, ethnicity, employment status, area deprivation education, and ever had a same sex relationship (model 2) and health measures (physical morbidities, BMI, ever seen GP for depression, psychotropic medication, alcohol consumption and smoking status) (model 3). Adjusting for loneliness (model 4) or emotional support (model 5) only led to a slight attenuation of associations, and in the final fully adjusted model (model 6) both living alone (Hazard Ratio (HR) 2.02, 95% CI 1.40 to 293) and living with a person who was not a partner (HR 1.72, 95% CI 1.03 to 2.88) were associated with death by suicide.

**Table 2:**
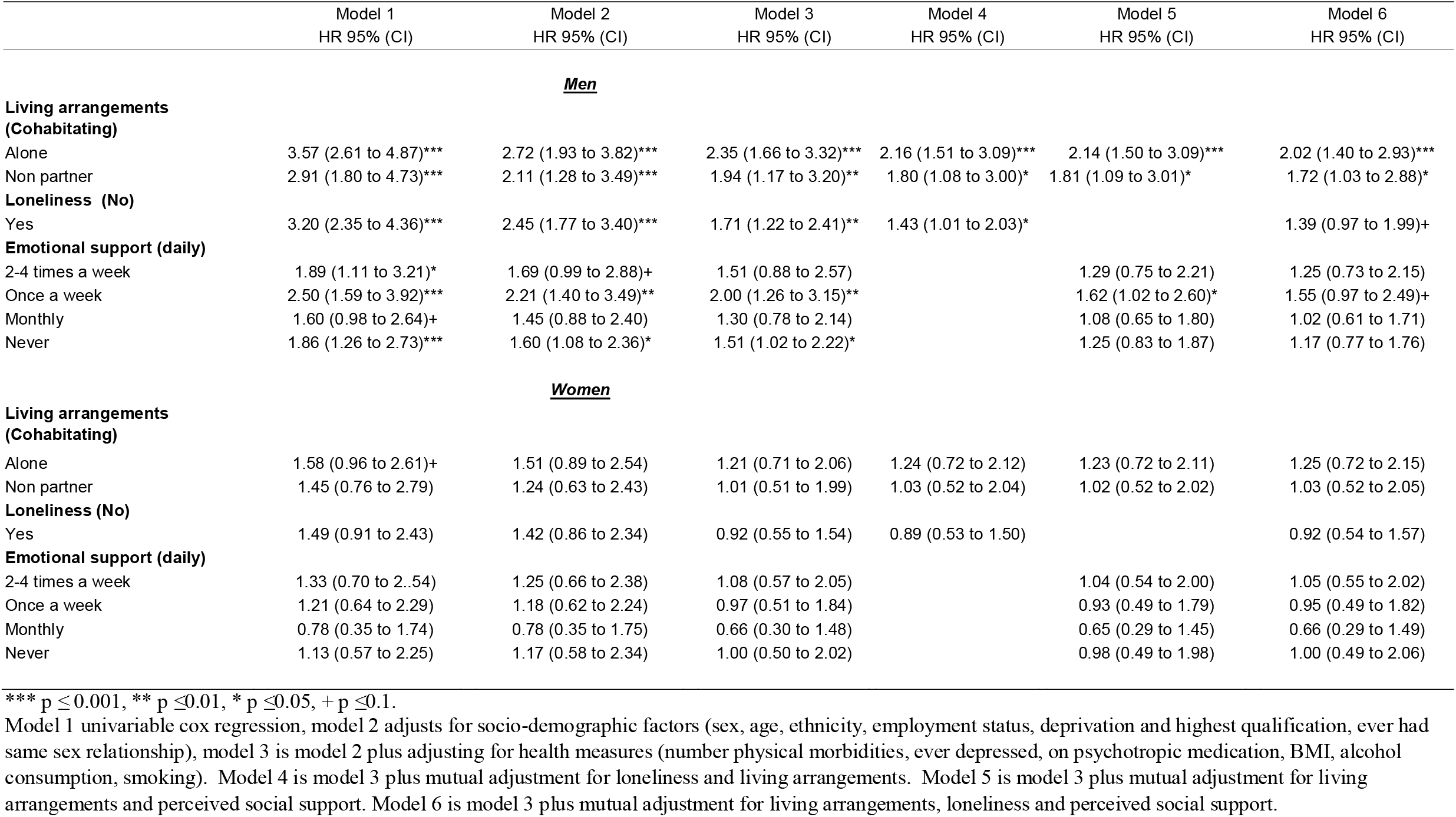
Death by suicide hazard ratios for loneliness, living arrangements and perceived social support, in unadjusted and adjusted models for men and women in the UK Biobank study

|  | Model 1<br>HR 95% (CI) | Model 2<br>HR 95% (CI) | Model 3<br>HR 95% (CI) | Model 4<br>HR 95% (CI) | Model 5<br>HR 95% (CI) | Model 6<br>HR 95% (CI) |
| --- | --- | --- | --- | --- | --- | --- |
| **<u>Men</u>** |  |  |  |  |  |  |
| **Living arrangements (Cohabiting)** |  |  |  |  |  |  |
| Alone | 3.57 (2.61 to 4.87)*** | 2.72 (1.93 to 3.82)*** | 2.35 (1.66 to 3.32)*** | 2.16 (1.51 to 3.09)*** | 2.14 (1.50 to 3.09)*** | 2.02 (1.40 to 2.93)*** |
| Non partner | 2.91 (1.80 to 4.73)*** | 2.11 (1.28 to 3.49)*** | 1.94 (1.17 to 3.20)** | 1.80 (1.08 to 3.00)* | 1.81 (1.09 to 3.01)* | 1.72 (1.03 to 2.88)* |
| **Loneliness (No)** |  |  |  |  |  |  |
| Yes | 3.20 (2.35 to 4.36)*** | 2.45 (1.77 to 3.40)*** | 1.71 (1.22 to 2.41)** | 1.43 (1.01 to 2.03)* |  | 1.39 (0.97 to 1.99)+ |
| **Emotional support (daily)** |  |  |  |  |  |  |
| 2-4 times a week | 1.89 (1.11 to 3.21)* | 1.69 (0.99 to 2.88)+ | 1.51 (0.88 to 2.57) |  | 1.29 (0.75 to 2.21) | 1.25 (0.73 to 2.15) |
| Once a week | 2.50 (1.59 to 3.92)*** | 2.21 (1.40 to 3.49)** | 2.00 (1.26 to 3.15)** |  | 1.62 (1.02 to 2.60)* | 1.55 (0.97 to 2.49)+ |
| Monthly | 1.60 (0.98 to 2.64)+ | 1.45 (0.88 to 2.40) | 1.30 (0.78 to 2.14) |  | 1.08 (0.65 to 1.80) | 1.02 (0.61 to 1.71) |
| Never | 1.86 (1.26 to 2.73)*** | 1.60 (1.08 to 2.36)* | 1.51 (1.02 to 2.22)* |  | 1.25 (0.83 to 1.87) | 1.17 (0.77 to 1.76) |
| **<u>Women</u>** |  |  |  |  |  |  |
| **Living arrangements (Cohabiting)** |  |  |  |  |  |  |
| Alone | 1.58 (0.96 to 2.61)+ | 1.51 (0.89 to 2.54) | 1.21 (0.71 to 2.06) | 1.24 (0.72 to 2.12) | 1.23 (0.72 to 2.11) | 1.25 (0.72 to 2.15) |
| Non partner | 1.45 (0.76 to 2.79) | 1.24 (0.63 to 2.43) | 1.01 (0.51 to 1.99) | 1.03 (0.52 to 2.04) | 1.02 (0.52 to 2.02) | 1.03 (0.52 to 2.05) |
| **Loneliness (No)** |  |  |  |  |  |  |
| Yes | 1.49 (0.91 to 2.43) | 1.42 (0.86 to 2.34) | 0.92 (0.55 to 1.54) | 0.89 (0.53 to 1.50) |  | 0.92 (0.54 to 1.57) |
| **Emotional support (daily)** |  |  |  |  |  |  |
| 2-4 times a week | 1.33 (0.70 to 2.54) | 1.25 (0.66 to 2.38) | 1.08 (0.57 to 2.05) |  | 1.04 (0.54 to 2.00) | 1.05 (0.55 to 2.02) |
| Once a week | 1.21 (0.64 to 2.29) | 1.18 (0.62 to 2.24) | 0.97 (0.51 to 1.84) |  | 0.93 (0.49 to 1.79) | 0.95 (0.49 to 1.82) |
| Monthly | 0.78 (0.35 to 1.74) | 0.78 (0.35 to 1.75) | 0.66 (0.30 to 1.48) |  | 0.65 (0.29 to 1.45) | 0.66 (0.29 to 1.49) |
| Never | 1.13 (0.57 to 2.25) | 1.17 (0.58 to 2.34) | 1.00 (0.50 to 2.02) |  | 0.98 (0.49 to 1.98) | 1.00 (0.49 to 2.06) |

\*\*\* $p \leq 0.001$ , \*\* $p \leq 0.01$ , \* $p \leq 0.05$ , + $p \leq 0.1$ .
Model 1 univariable cox regression, model 2 adjusts for socio-demographic factors (sex, age, ethnicity, employment status, deprivation and highest qualification, ever had same sex relationship), model 3 is model 2 plus adjusting for health measures (number physical morbidities, ever depressed, on psychotropic medication, BMI, alcohol consumption, smoking). Model 4 is model 3 plus mutual adjustment for loneliness and living arrangements. Model 5 is model 3 plus mutual adjustment for living arrangements and perceived social support. Model 6 is model 3 plus mutual adjustment for living arrangements, loneliness and perceived social support.

The relationship between loneliness and low levels of emotional support and death by suicide, although relatively strong in unadjusted models (model 1), fell after adjustment for sociodemographic factors and health. Once living arrangements were accounted for, loneliness (model 4), and lower levels of emotional support (model 5) were only modestly associated with death by suicide. In contrast, for women, there was little evidence of any association between death by suicide and living arrangements, loneliness or emotional support.

Finally, in fully adjusted models, we conducted interaction tests to assess whether the relationship between living arrangements and death by suicide was modified by loneliness or emotional support. For men, a Wald test indicated a significant interaction (p=0.002) between living arrangements and loneliness, presented in Figure 1a. Men who often experienced loneliness or those who were not lonely and living alone, or with a non-partner only, had three times the risk of dying by suicide compared to those who cohabit and are not lonely.

**Figure 1:**
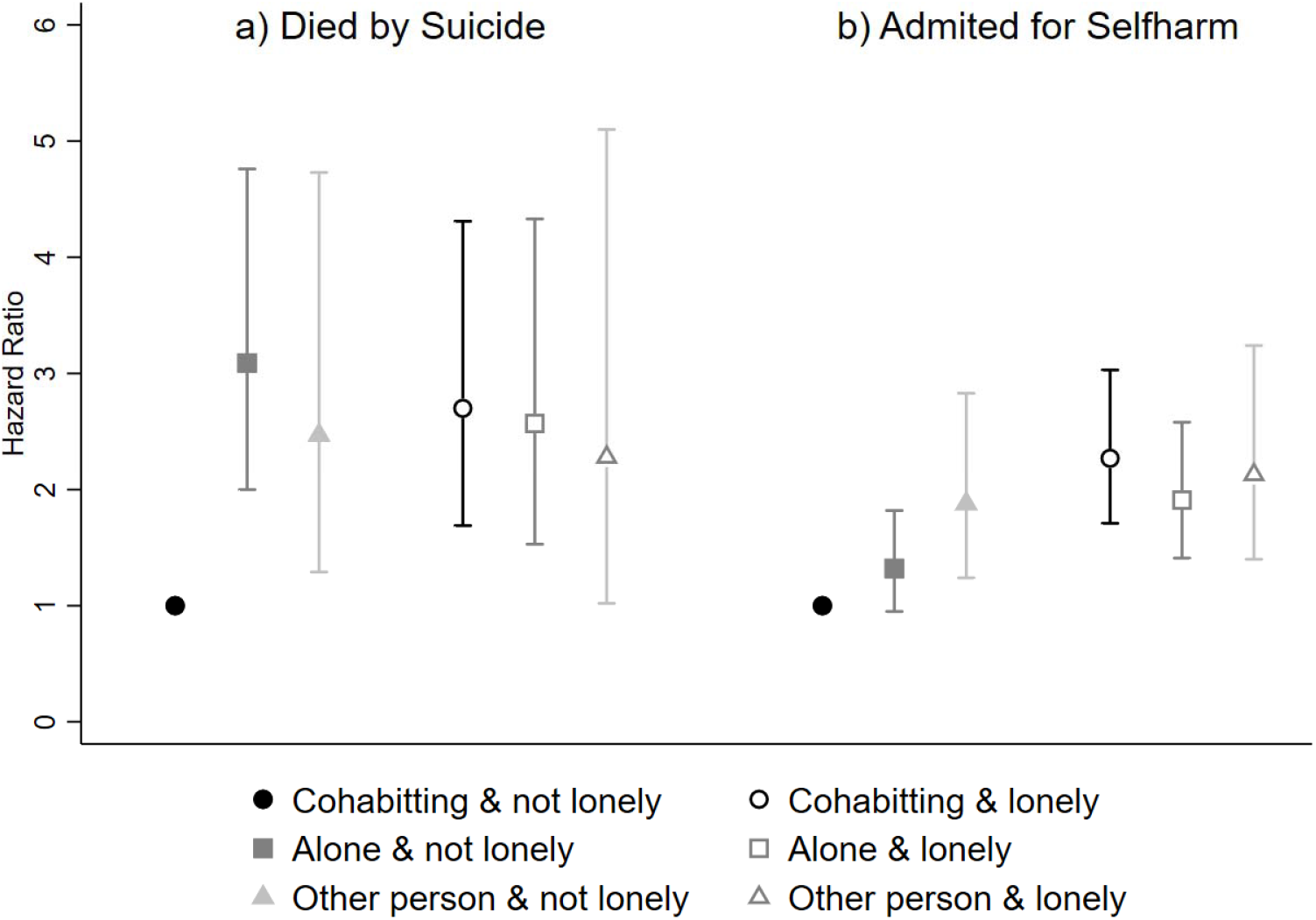
The interactions between living arrangements and loneliness in the prediction of deaths by suicide and hospital admissions for self-harm for men in the UK Biobank Study.

### Hospital admissions for self-harm

The results for the Royston Parmar models for associations between hospital admissions for self-harm and living arrangements, loneliness and perceived emotional support are shown in table 3.

**Table 3:** Admissions to hospital for self-harm hazard ratios for loneliness, living arrangements and perceived social support, in unadjusted and adjusted models for men and women in the UK Biobank study

|  | Model 1<br>HR 95% (CI) | Model 2<br>HR 95% (CI) | Model 3<br>HR 95% (CI) | Model 4<br>HR 95% (CI) | Model 5<br>HR 95% (CI) | Model 6<br>HR 95% (CI) |
| --- | --- | --- | --- | --- | --- | --- |
| **<u>Men</u>** |  |  |  |  |  |  |
| **Living arrangements (Cohabiting)** |  |  |  |  |  |  |
| Alone | 3.14 (2.57 to 3.84)*** | 1.67 (1.34 to 2.09)*** | 1.37 (1.09 to 1.72)** | 1.17 (0.93 to 1.48) | 1.17 (0.93 to 1.48) | 1.06 (0.83 to 1.34) |
| Non partner | 3.56 (2.70 to 4.70)*** | 1.77 (1.32 to 2.38)*** | 1.64 (1.23 to 2.20)*** | 1.44 (1.07 to 1.94)* | 1.44 (1.10 to 1.95)* | 1.33 (0.98 to 1.79)+ |
| **Loneliness (No)** |  |  |  |  |  |  |
| Yes | 5.32 (4.43 to 6.40)*** | 3.13 (2.57 to 3.82)*** | 2.01 (1.63 to 2.47)*** | 1.91 (1.55 to 2.37)*** |  | 1.74 (1.40 to 2.16)*** |
| Per log day X Loneliness | 0.97 (0.93 to 1.01)+ | 0.97 (0.93 to 1.01)+ | 0.97 (0.93 to 1.01)+ | 0.97 (0.93 to 1.01)+ |  | 0.97 (0.93 to 1.01)+ |
| **Emotional support (daily)** |  |  |  |  |  |  |
| 2-4 times a week | 1.95 (1.37 to 2.77)*** | 1.68 (1.18 to 2.40)** | 1.47 (1.03 to 2.10)* |  | 1.41 (0.99 to 2.02)+ | 1.34 (0.94 to 1.92)+ |
| Once a week | 2.46 (1.82 to 3.32)*** | 2.01 (1.48 to 2.72)*** | 1.79 (1.32 to 2.42)*** |  | 1.69 (1.24 to 2.32)*** | 1.56 (1.14 to 2.14)** |
| Monthly | 2.13 (1.58 to 2.88)*** | 1.84 (1.34 to 2.49)*** | 1.60 (1.18 to 2.17)** |  | 1.53 (1.13 to 2.09)** | 1.39 (1.02 to 1.90)* |
| Never | 2.98 (2.38 to 3.74)*** | 2.16 (1.72 to 2.72)*** | 1.95 (1.54 to 2.46)*** |  | 1.86 (1.46 to 2.36)*** | 1.65 (1.29 to 2.10)*** |
| **<u>Women</u>** |  |  |  |  |  |  |
| **Living arrangements (Cohabiting)** |  |  |  |  |  |  |
| Alone | 1.75 (1.44 to 2.11)*** | 1.67 (1.37 to 2.04)*** | 1.18 (0.96 to 1.44) | 1.04 (0.85 to 1.28) | 1.15 (0.93 to 1.41) | 1.04 (0.85 to 1.28) |
| Non partner | 2.00 (1.60 to 2.50)*** | 1.21 (0.96 to 1.53) | 0.94 (0.74 to 1.19) | 0.85 (0.67 to 1.08) | 0.92 (0.73 to 1.17) | 0.85 (0.67 to 1.08) |
| **Loneliness (No)** |  |  |  |  |  |  |
| Yes | 4.35 (3.60 to 5.13)*** | 3.25 (2.74 to 3.85)*** | 1.92 (1.61 to 2.29)*** | 1.93 (1.61 to 2.31)*** |  | 1.89 (1.57 to 2.28)*** |
| Per log day X Loneliness | 0.95 (0.93 to 0.98)*** | 0.95 (0.93 to 0.98)*** | 0.95 (0.93 to 0.98)*** | 0.95 (0.93 to 0.98)*** |  | 0.95 (0.93 to 0.98)*** |
| **Emotional support (daily)** |  |  |  |  |  |  |
| 2-4 times a week | 1.21 (0.92 to 1.60) | 1.18 (0.90 to 1.56) | 0.99 (0.75 to 1.31) |  | 0.98 (0.74 to 1.30) | 0.91 (0.69 to 1.21) |
| Once a week | 1.52 (1.19 to 1.94)*** | 1.50 (1.17 to 1.92)*** | 1.20 (0.94 to 1.54) |  | 1.18 (0.92 to 1.52) | 1.05 (0.82 to 1.35) |
| Monthly | 1.26 (0.98 to 1.66)+ | 1.27 (0.97 to 1.67)+ | 1.03 (0.78 to 1.35) |  | 1.02 (0.78 to 1.34) | 0.88 (0.67 to 1.16) |
| Never | 2.20 (1.76 to 2.76)*** | 1.94 (1.54 to 2.43)*** | 1.50 (1.19 to 1.89)*** |  | 1.49 (1.18 to 1.88)*** | 1.24 (0.98 to 1.58)+ |

\*\*\* $p \leq 0.001$ , \*\* $p \leq 0.01$ , \* $p \leq 0.05$ , + $p \leq 0.1$ .
Model 1 Univariable Royston Parmar parametric Hazard models, model 2 adjusts for socio-demographic factors (sex, age, ethnicity, employment status, deprivation and highest qualification, ever had same sex relationship). Model 3 is model 2 plus adjusting for health measures (number physical morbidities, ever depressed, on psychotropic medication, BMI, alcohol consumption, smoking). Model 4 is model 3 plus mutual adjustment for loneliness and living arrangements. Model 5 is model 3 plus mutual adjustment for living arrangements and perceived social support. Model 6 is model 3 plus mutual adjustment for living arrangements, loneliness and perceived social support.

For men, all of living arrangements, loneliness and perceived emotional support were associated with hospital admissions for self-harm in unadjusted analyses. The strength of these associations was reduced after adjusting for sociodemographic characteristics (model 2) and health measures (model 3). Both loneliness (model 4) and lower levels of emotional support (model 5) explained part of the relationship between living alone and self-harm. In the final model, there was no evidence of an association between living alone and self-harm. In contrast, both lower levels of emotional support and loneliness were associated with increased risk of hospitalization for self-harm.

Women differed from men in that the living arrangements categories had weaker relationships with self-harm within the unadjusted model (model 1). Furthermore, the associations for women were explained by health (model 3). For women, loneliness (model 4) and lower levels of perceived emotional support (model 5) were associated with increased risk of self-harm, independent of other factors (with the caveat that the relationship between lack of emotional supports and self-harm could be explained by loneliness).

For men, adjusting for all confounders, we found a significant (p=0.023) interaction indicating that loneliness modified the relationship between living arrangements and hospitalization for self-harm. Overall, loneliness removed any protective associations of cohabitation over living alone, such that men who were often lonely had similarly increased HRs of hospitalization of around 2, irrespective of their living arrangements. In contrast, among men who did not report loneliness, living alone was associated with a modest increase in risk of hospitalization for self-harm (HR 1.32, 95% CI 0.95 to 1.82) and a greater increase was found for those living only with non-partners (HR 1.88, 95% CI 1.24 to 2.83) (see Figure 1b).

## DISCUSSION

Our goal was to investigate the association between living arrangements, loneliness, perceived emotional support and subsequent risk of suicidal behaviours, within a large general population cohort in middle-age.

For men, given that both living alone and living with a non-partner were both associated with an increased risk of death by suicide, it is possible that having a partner is protective against death by suicide. Subjective loneliness and perceived emotional support had modest relationships with death by suicide and these variables explained little of the relationship between living alone and death by suicide. For women, none of living arrangements, loneliness, nor emotional support were associated with death by suicide.

For both men and women, loneliness and emotional support were associated with increased risk of hospitalization for self-harm. However, associations between living arrangements and self-harm were more limited, being explained by health for women, and health, loneliness and emotional support for men.

Our study has some important novel findings. While loneliness has been linked to suicidal ideation and attempts (Stickley and Koyanagi, 2016), including within case-control studies (Sinclair et al., 2005), a recent systematic review (Solmi et al., 2020) suggests that our study is the first longitudinal study of its kind to investigate the relationship between loneliness and deaths by suicide. The absence of prior research is not a surprise given the methodological challenges of studying such rare outcomes as suicide (Van Orden et al., 2010).

Our finding that deaths by suicide and hospital admissions for self-harm for men were associated with living arrangements, is consistent with the literature for marital status that finds that married or cohabiting people have lower risks of suicide compared to single people (Conejero et al., 2016; Frisch and Simonsen, 2013). Our study adds to this literature by showing that for men actually living with a partner (not just from not living alone) appears to be associated with reduced risk of death by suicide and hospital admissions for self-harm. Demographic factors, such as the older age of the UK Biobank sample, could explain why we did not find any associations between living arrangements and suicide and self-harm for women. Kyung-Sook et al. (2018) found associations between marital status and suicide only among younger women.

An original contribution of our study with respect to living arrangements is that most studies of suicidal behaviour focus on the concept that it is *living alone* that is harmful (Turecki and Brent, 2016). However, our results indicate that both men who lived alone *and with non-partners* were at increased risk of dying by suicide and self-harm. Apart from Frisch and Simonsen’s (2013) study, which indicated that people living in households with more than nine people had increased risk of suicide, the focus has generally been on living alone as a risk factor, rather than other relationships. However, this is to some extent consistent with a wider literature which has found that those living alone and those living with people other than a partner are more likely to have anxiety or depressive disorders (Joutsenniemi et al., 2006).

Another original contribution with respect to living arrangements is that our results are the first to indicate that the associations between living alone and self-harm might be explained by loneliness and emotional support. However, this does not appear to be the case for deaths by suicide. Such findings, particularly if replicated elsewhere, may have implications for theories that try to explain the psychological and social antecedents of suicidal behaviour.

### Implications

As noted above, the rarity of death by suicide as an outcome has made it very difficult to investigate some theorised risks for death by suicide, as well as other relatively infrequent outcomes such as hospitalization for self-harm (Klonsky et al., 2016; Van Orden et al., 2010). Many of the theories proposed to explain the development of suicidal behaviour are based on studies investigating risk factors for other forms of suicidal behaviour, such as self-reported suicidal ideation or attempts. Clearly, the assumption that the risk factors for all types of suicidal thoughts and behaviours will be the same is problematic (DeJong et al., 2010; Klonsky et al., 2016). From a public health perspective, it is important to identify potentially causal and modifiable factors that may differ between death by suicide and other suicidal behaviours.

This study clearly shows that, for men, living arrangements, loneliness and emotional support are important risk factors for both death by suicide and hospitalization for self-harm. For women, loneliness and lack of emotional support are important risk factors for hospitalization for self-harm. Our results are less certain with respect to the role that living arrangements, loneliness and emotional support might play in deaths by suicide for women. Given the large differences in rates of death by suicide between men and women (Scourfield and Evans, 2015) and systematic review findings that the relationship between marital status and suicide is moderated by gender (Kyung-Sook et al., 2018), *a priori* we decided to analyse the results separately by gender. The large gender difference in death by suicide rates indicates that either risk factors’ associations or their prevalence differ by gender. From our stratified analyses, we find much more modest associations (around half the strength of equivalent associations for men) between living arrangements and loneliness and death by suicide in unadjusted models for women, and these associations are largely explained by socioeconomic factors, health and mental health at baseline. The confidence intervals for the sample do not completely rule out the possibility of there being associations between death by suicide and living arrangements, loneliness and emotional support for women within the general population. To carry out improved analyses for women would require much larger samples, such as national-level UK census linked to hospital and mortality records. Even then, only some aspects of our analyses could be replicated as these administrative data lack many important variables, particularly loneliness.

Our findings on the relationship between death by suicide and living arrangements, loneliness and emotional support for men suggest that these are not simply distal risk factors: the associations persist after adjusting for measures of physical and mental health. With respect to death by suicide, the protective associations of living with a partner are particularly important. Cohabiting with a partner appears to be associated with protection against death by suicide even after adjusting for socioeconomic and demographic factors, physical health, mental health, loneliness and emotional support. This could be the case for a number of reasons but operationalizing loneliness or emotional support using single item measures is unlikely to be the explanation. The loneliness and emotional support measures we used were strongly associated with self-harm. An alternative methodological explanation is that men who die by suicide may be less willing to seek treatment for poor mental health and that the risk of suicide is greater among those who never see a GP, compared to those who see a GP once a year (Windfuhr et al., 2016), hence there may be residual confounding due to poor health. However, that would not in itself explain why men who are not living with a partner have increased risk of poor mental health, that in turn leads to death by suicide.

There are also theoretical reasons that could explain why having a partner is protective against death by suicide. One possibility is that the benefits of having a partner may be linked to men’s sense of masculinity and self-image (Scourfield et al., 2012), rather than emotional support or companionship. Given that living with a non-partner does not appear to be associated with any protective effects, and the removal of the protective associations of having a partner among people who are lonely, some of the risk might be due to the concept of “perceived burdensomeness” from the Interpersonal Theory of Suicide (Van Orden et al., 2010). Components of perceived burdensomeness, which include self-hatred and feeling so flawed that one becomes a liability to others, could be theorized as being active drivers for death by suicide. Perceived burdensomeness could arise in situations in which living arrangements suggested a dysfunctional relationship (such as being lonely while cohabiting) or living in situations in which men are unable to fulfil traditional male roles that require having a partner (Scourfield and Evans, 2015). It is possible that perceived burdensomeness may not just be a driver of suicidal behaviour in general, but also a driver towards more lethal self-harm behaviours.

For hospitalization for self-harm, in contrast to death by suicide, loneliness and emotional support appear to be the more important than living arrangements. Associations exist after adjusting for sociodemographic and mental and physical health factors, and for men loneliness and emotional wellbeing explain most of the relationship between living alone and hospitalization for self-harm. This is consistent with the idea that loneliness and lack of emotional support are mechanisms through which living alone, or at least without a partner, could increase risk of suicide. This is also consistent with the concept of ‘thwarted belongingness’ from the Interpersonal Theory of Suicide, which suggests that loneliness alongside the absence of reciprocal caring relationships can lead to self-destructive behaviours (Stanley et al., 2016; Van Orden et al., 2010). However, given that in the presence of loneliness any protective associations of cohabitation are removed for men, it also raise questions about the extent to which a lonely person can also feel that they do or do not have a reciprocal caring relationship.

Our findings are consistent with some differences in the risks for different types of suicidal behaviour (DeJong et al., 2010; Klonsky et al., 2016). One possibility is that while thwarted belongingness and perceived burdensomeness both drive suicidal behaviour, the latter more strongly drives individuals towards more lethal methods. However, our results may also be consistent with other theories for suicidality. An alternative is that loneliness could be considered a form of emotional dysregulation. Emotional dysregulation theory proposes that while emotional dysregulation is an important factor in self-harm, some aspects of emotional dysregulation may be protective against something that is as daunting and as fearful as lethal self-harm (Stanley et al., 2016). It should be noted that lack of emotional support was more strongly associated with self-harm than it was associated with death by suicide. Given that emotionally supportive relationships can improve emotional regulation (Overall and Simpson, 2013), this would also be consistent with emotional dysregulation theory.

### Strengths and limitations

The key strength of this study is that UK Biobank had a baseline sample of more than 500,000 people. This very large sample provided an opportunity to study death by suicide and hospitalization for self-harm, which are both rare outcomes. However, UK Biobank data do have some potential limitations. The recruitment of such a large sample is only justifiable if it collects data on a broad range of topics, many of which are necessarily operationalised using single item questions. The use of single item measures for loneliness, which was a simple dichotomous measure, and perceived emotional support might be considered a weakness of our study. They are items drawn from longer scales and have not been validated as single items. The extent of loneliness may be underreported as there are negative connotations to being lonely and people may not always admit that they feel lonely (de Jong Gierveld, 1998). However, single-item measures are considered appropriate (Stickley and Koyanagi, 2016) and have been recommended for the study of loneliness (HM Government, 2018). Another limitation is that UK Biobank had an invitation response rate of only 5.5%, and, compared to the general population UK Biobank is less economically deprived with some evidence of a healthy volunteer selection bias (Fry et al., 2017). A heathy volunteer selection bias may explain why the yearly death by suicide rates for both men (8.9 deaths per 100,000 per year) for women (3.5 deaths per 100,000 per year) in UK Biobank is lower than death rate for suicides in the UK (Office for National Statistics, 2019).

Our study uses observational data with the inherent limitations for inferring causality. We have adjusted for a broad range of potential confounding variables at baseline and we had follow-up data on deaths by suicide and hospitalisation for self-harm. However, our baseline measures were only recorded at a single time point and potentially participants’ circumstances could have changed over time. In addition, with measurements recorded at only one time point, it is impossible to determine the causal direction between measures conclusively. It is likely that the relationship between the social connection measures and baseline health is bidirectional. However, given that the peak age of onset for major depressive disorder is early adulthood (Myrna M. Weissman et al., 2016), we decided to focus on a somewhat conservative approach, prioritising relationships presented in models that have adjusted for both mental health and sociodemographic measures. The mental health measures that are available for all UK Biobank participants at baseline were: self-report of ever seeing a GP for nerves, anxiety, and depression; and receipt of psychotropic medication. This may underestimate depression, which might confound the associations found in the study. In addition, the frequency of alcohol consumption measure will not fully capture substance abuse. There are other potential confounding measures for which data is unavailable in UK Biobank including personality disorders, conflict and stressful life events. We were also limited in our analyses by only having self-harm hospital admission data for England (these data were not available for Wales or Scotland). It is also the case that many who self-harm do not seek help from services or, when they do, are not admitted but are reviewed as out-patients (Gunnell et al., 2005).

Finally, the target sample of this population was those aged 40 to 70 living in the United Kingdom, and the sample contained only a handful of people outside this age range. These results may not be generalizable to other age groups or to other cultures where attitudes to suicide, or other societal level risk factors for suicide, such as the availability of fire arms, may be very different.

## CONCLUSIONS

This study raises several questions for future exploration. Our results suggest that addressing loneliness in the general population may reduce the risk of self-harm but, for death by suicide, there is a much more complex (and likely sex-specific) relationship between loneliness, living arrangements and perceived emotional support. Overall, this work demonstrates that for men (but not for women) living alone or with a non-partner is associated with increased risk of suicide, a finding not explained by perceived loneliness. It appears likely that loneliness may be more important as a risk factor for self-harm than for suicide. These findings may reflect differences in the theoretical pathways for death by suicide and self-harm.

## Data Availability

UK Biobank is an open access resource available to verified researchers upon application.

http://www.ukbiobank.ac.uk/

## AUTHOR STATEMENTS

### Contributors

RJS, DJS, BC and AJ developed the research question. NG, JW, BC, RJS, AJ, DML, RP assisted with deriving and coding the variables. RJS carried out the analysis. BC, DML and DM provided advice on statistical methodology. AJ and CO carried out the literature review. RJS, DJS, AJ and CO drafted the manuscript. All authors edited and agreed the final manuscript.

### Funding sources

This project was supported by two MRC Mental Health Data Pathfinder Awards [grant numbers MC_PC_17217, MC_PC_17211].

## Acknowledgements

We thank all participants in the UK Biobank study. The project was completed using UK Biobank project code 6553 (PI: Smith). UK Biobank was established by the Wellcome Trust, Medical Research Council, Department of Health, Scottish Government and Northwest Regional Development Agency. UK Biobank has also had funding from the Welsh Assembly Government and the British Heart Foundation. Data collection was funded by UK Biobank.

## Notes

**Declarations of interest**: none.

### Competing Interest Statement

The authors have declared no competing interest.

### Funding Statement

This project was supported by two MRC Mental Health Data Pathfinder Awards (D.J.S., grant number MC_PC_17217),(A.J., grant number MC_PC_17211).

### Author Declarations

All relevant ethical guidelines have been followed and any necessary IRB and/or ethics committee approvals have been obtained.

Any clinical trials involved have been registered with an ICMJE-approved registry such as ClinicalTrials.gov and the trial ID is included in the manuscript.

